# Transfer Language Space with Similar Domain Adaptation: A Case Study with Hepatocellular Carcinoma

**DOI:** 10.1101/2020.08.26.20182659

**Authors:** Patricia Balthazar, Scott Jeffery Lee, Terry Desser, Daniel L. Rubin, Judy Wawira Gichoya, Imon Banerjee

## Abstract

Transfer learning is a common practice in image classification with deep learning where the available data is often limited for training a complex model with millions of parameters. However, transferring language models requires special attention since cross-domain vocabularies (e.g. between news articles and radiology reports) do not always overlap as the pixel intensity range overlaps mostly for images. We present a concept of similar domain adaptation where we transfer an inter-institutional language model between two different modalities (ultrasound to MRI) to capture liver abnormalities. Our experiments show that such transfer is more effective for performing shared targeted task than generic language space transfer. We use MRI screening exam reports for hepatocellular carcinoma as the use-case and apply the transfer language space strategy to automatically label thousands of imaging exams.

## Introduction

Hepatocellular carcinoma (HCC) is the most common primary liver malignancy and a leading cause of cancer death worldwide^1^. Imaging is critical for HCC screening and includes ultrasound and multiphase cross-sectional imaging using CT and MRI^2^. Unlike most cancers, imaging can be used to establish the definitive diagnosis of HCC noninvasively, and treatment (including major surgical options such as hepatic resection and liver transplant) is usually initiated without confirmatory biopsy^3^. HCC surveillance programs aim to provide timely diagnosis and detect early preclinical stage of disease where cure is possible with locoregional therapies or liver transplantation, which ultimately reduces mortality^4^.

Although ultrasonography (US) is identified as the most cost-effective tool for HCC screening, MRI is more sensitive and has a specificity of > 90 percent. Thus, in cirrhotic and other high-risk patients (e.g., Asian and African American origin, obese, diabetic, alcohol addicted, hepatitis infection) MRI/CT are frequently used for screening. Patients frequently receive screening studies across multiple modalities based on various institution protocols, resulting in a mix of US, CT and MRI images when reviewing a longitudinal patient record. Several radiology societies have developed structured radiology report templates and standardized coding systems to promote secondary use of the large amounts of unstructured data that exists in radiology departments^5^. One such effort is the Liver Imaging Reporting and Data System (LI-RADS)^4^ developed by the American College of Radiology (ACR) and adopted as the standard coding system for HCC screening in 2018. Despite these efforts, adoption of structured reports remains low^6^, and simple tasks like differentiating between benign and malignant exams from a large number of screening cases, requires hundreds of hours of manual review by the radiologist. In our institution, which is a large multi-site academic center, structured reports coded with LI-RADS were adopted in 2018, representing only a small fraction of available HCC screening data.

Traditional natural language processing (NLP) methods (rule-based, dictionary-based, supervised learning) have been successfully applied to radiology reports for extracting information or performing automatic report classification^7,8^. NLP techniques have also been explored for information extraction from various liver studies for colorectal adenoma^9^, inflammatory bowel disease^10^, fatty liver disease^11^, aortic aneurysm^8^, and liver steatosis^12^. However, a major limitation of traditional NLP techniques is the requirement of large-scale human-labeled data or explicit hand-written linguistic rules that limit both scalability and generalizability of the system. Moreover, the use of experts to perform required labeling is expensive, and hence medical data annotation is performed on a small cohort of studies. We previously developed a word embedding-based NLP pipeline^13^ for inferring LI-RADS coding by considering only the liver findings written in the narrative US reports where only a minimal amount of hand-curated data is required for training.

We have expanded our previous work on word embedding for US LI-RADS ^13,14,15^ to a new institution and a new modality (MRI) coding to build a system that will automatically label a large number of imaging exams to generate a large-scale annotated MRI dataset (benign and malignant). Our approach leverages the benefits of unsupervised learning (word2vec^16^) along with expert-knowledge to tackle the major challenges of information extraction from radiology texts, which include ambiguity of free text narrative, lexical variations, use of ungrammatical and telegraphic phases, arbitrary ordering of words, and frequent appearance of abbreviations and acronyms. This method transforms the free-text radiology reports into dense vectors that help unlock rich source of information for computerized parsing. Distributional semantics models (word2vec) are usually trained on more than millions of similar articles/reports. In comparison, we had a limited number of MRI reports for HCC screening available at our institution. In order to handle the data scarcity, we use *transfer learning with similar domain adaptation* where we fine-tune a language model (LM) which was originally trained on HCC screening US reports, and subsequently applied on the MRI reports. Our hypothesis is that modality shifted similar domain adaptation, even if fine-tuned on a limited data set, is able to capture semantics of radiology better than other LMs trained on completely different domains. This hypothesis is built on the premise that radiologic term representations learned for one task can be directly useful for another task if the domains are analogous. Using this approach allows for curation of large scale datasets for machine learning combining both image/pixel and text information. In this paper, we prove our hypothesis by fine-tuning a language model (LM) trained on US reports from Stanford Health Care (SHC) to infer the labels from MR studies (n = 10,000, retrospective) performed at Emory University Healthcare (EUH). To demonstrate the benefit of similar domain adaptation, we compare the performance of the fine-tuned LM (from EUH) with the directly transferred US language model from SHC and a second comparison of the EUH LM with the LM initially trained on 3 million Google news articles.

## Method

Figure 1 presents our experimentation pipeline where we are using 3 different cohorts to train the LMs-(1) SHC ultrasound reports cohort of HCC screening to initially train a LM; (2) Google news articles to initially train a LM; (3) EUH MRI reports of HCC screening to fine-tuned the LMs. On top of the LM, we leveraged the semi-structured EUH MRI reports (recent) to train the discriminative classifier for identifying the labels and used both semi-structured and unstructured (free-text) EUH MRI reports to validate the performance. In order to explain complexity of the task, we present a sample structured and unstructured MRI report in Figure 2.

**Figure 1:**
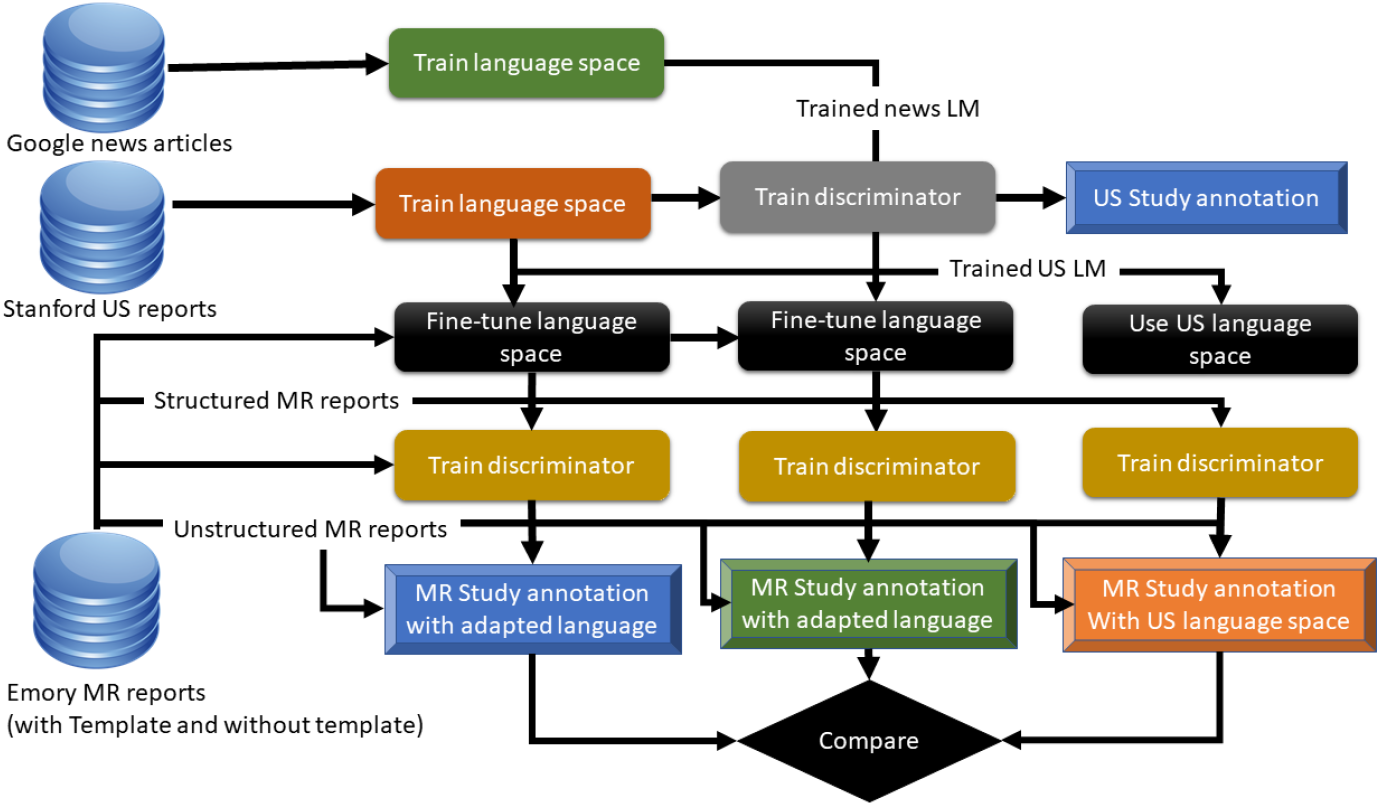
Flow diagram of language model transfer from US to MRI and GoogleNews to MRI

**Figure 2:**
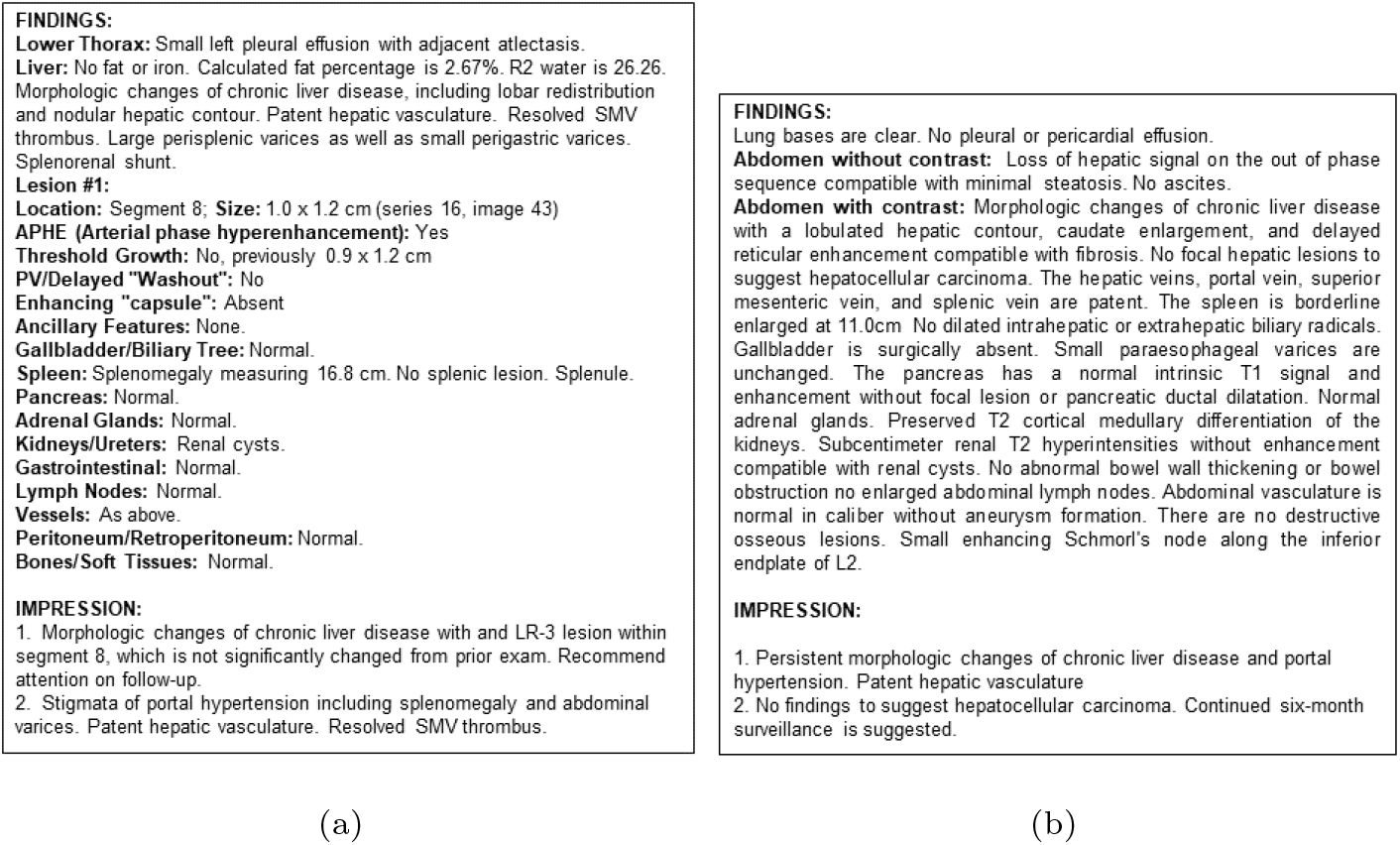
Sample MR reports: (a) Sample with LI-RADS structured template, (b) Sample free-text report

### Dataset

*Stanford US dataset:* With the approval of Stanford Institutional Review Board (IRB), we collected all the free-text radiology reports of abdominal ultrasound examinations performed at Stanford Hospital between August 2007 to December 2017. In total, there were 92,848 US reports collected over 10 years, with an average 9,000 US exams performed every year. Among them, 13,860 exams were performed for HCC screening. A total 1,744 abdominal US studies were coded with the US LI-RADS reporting format.

*EUH MRI dataset:* With the approval of Emory University IRB, we retrieved 10,018 MRI exams performed between 2010 – 2019 at EUH for HCC screening. Among these, only 548 studies were reported using the LI-RADS structured template where a unique LI-RADS score was reported in the impression section (Figure 2.a.). From the LI-RADS coded reports, 99% were malignant cases (LR score >2) since benign cases are often not coded with LI-RADS. 9,470 MRI abdomen exams were documented as free-text narratives where the final diagnosis was recorded without following any structured scoring schema (Figure 2.b.).

We developed simple regex expressions to extract the LI-RADS codes from the impression section of the EUH semi-structured MRI reports. To obtain a representative sample of benign cases (which represented 1% of the LI-RADS coded reports), two radiologists manually annotated 537 benign cases from the EUH MRI dataset. We selected benign cases from reports performed after 2018 in order to match the report structure of annotated malignant cases.

To generate a large MRI dataset of HCC screening, the aim of the study was to label the 9,470 MRI abdomen free-text reports obtained between 2010 – 2017 without the LI-RADS formatted template. However, ground truth labels were missing for these reports since LI-RADS coding was not adopted before 2018 in our institution. To generate the test set, two radiology residents supervised by a board-certified radiologist independently assigned labels to a set of randomly selected 112 exams by analyzing only the free-text reports.

In Table 1, we present the synopsis of the Stanford US dataset and EUH MRI dataset according to report-level and word-level statistics which reflects a slight diversity between the style of reporting. For instance, the number of words in the reports ranged from 24 to 331 in the US dataset while for MR dataset in varies from 11 to 2116. Same observation holds for the number of sentences in the reports. It is also interesting to note here that 1790 common words between MR and US.

**Table 1:** Statistics of the cohorts before processing – Stanford US dataset and EUH MRI dataset.

|  |  |  |
| --- | --- | --- |
|  | Stanford US dataset | EUH MRI dataset |
| Cohort level |  |  |
| Number of unique words | 17194 | 19828 |
| Common words in two domains | 1790 |  |
| Report level |  |  |
| Average number of words (+/- std) | 167 (+/- 39) | 197 (+/-47) |
| Average number of sentences (+/- std) | 27 (+/-7) | 32 ( (+/-8) |

### Report pre-processing

*Segmentation* We design a python-based liver section segmentation module that works with both MRI/CT and US reports. The model uses a combination of regular expressions and dictionary based sentence retrieval using anatomical vocabularies derived from Foundational Model of Anatomy (FMA)^17^ to extract only findings related to liver and its sub-regions from the whole report. The module maintains dependencies between anatomical entities (e.g. all the lesions found within the liver would be extracted even if they are not described in the same paragraph). This segmentation module has been manually validated on randomly selected 100 MRI and 100 US reports and obtained perfect accuracy for segmenting the liver section and liver related statements from both recent (more structured) and historic radiology reports. In order to perform a valid experiment from the LI-RADS formatted US and MRI reports, we excluded the Impression section of the reports since the final LI-RADS assessment category is often reported explicitly in the Impression. The Findings section includes only the imaging characteristics of the liver abnormalities; thus, it does not contain any clear-cut definition of the LI-RADS final assessment classes.

*Root term mapping* We used a dictionary-style controlled-term mapping technique, where publicly available CLEVER terminology was used to replace common analogies/synonyms to create more semantically structured texts. As shown in our previous studies^15,14,13^, this strategy helps to reduce the number of unique words in the vocabulary and optimizes the learning for the distributional semantics model. We focused on the terms that described family, progress, risk, negation, and punctuations, and normalized them using the formal terms derived from the terminology. For instance, ‘mother’, ‘brother’, ‘wife’ .. →‘FAMILY’, ‘no’, ‘absent’, ‘adequate to rule her out’ .. → ‘NEGEX’, ‘suspicion’, ‘probable’, ‘possible’ → ‘RISK’, ‘increase’, ‘invasive’, ‘diffuse’, .. →‘QUAL’.

*Text cleaning* We normalize the text by converting in to lowercase letters, removing general stop words (e.g. ‘a’, ‘an’, ‘are’, ‘be’, ‘by’, ‘has’, ‘he’, etc.), removing words with very low frequency (< 50). We also removed unwanted terms/phrases (e.g. medicolegal phrases such as “I have personally reviewed the images for this examination”); these words generally appear in all or very few reports, and are thus of little to no value in document classification. We used the Natural Language Tool Kit (NLTK) library^18^ for determining the stop-word list and discarded these words during report indexing. We also discarded radiologist, clinician, and patient-identifying details from the reports. Date and time stamps were replaced by ‘<datetoken>’ phase.

### Initial training with Stanford US reports

In our previous study^13^, we leveraged 92,848 US reports collected over 10 years at Stanford hospital to train a word2vec model to learn the similarity between various terms used by different radiologists for reporting the liver US findings across a large time window in an unsupervised way. The primary goal was to successfully map the vocabulary used before (old reports) and after LI-RADS template implementation (new reports). The word2vec model adopts distributional semantics to learn dense vector representations of all words in the pre-processed corpus by analyzing the context. We trained the model on a pre-processed corpus where we extracted the liver section and the model learnt the representation of 2000 unique words. The model demonstrated encouraging performance for learning a LI-RADS vocabulary (semantic dictionary) using word similarity clustering. The ensemble machine-learning model trained on top of the embedding was also able to infer the three LI-RADS final assessment categories from the free-text US reports and achieve performance comparable with human raters^13^.

### Fine-tuning with MRI reports

In the current study, given the limited number of MRI exams (10,018) available to train a language model from scratch, we transferred the language model trained on US reports from Stanford hospital to MRI corpus at EUH. Although both MRI and US LI-RADS exams were targeted to provide a standard diagnostic algorithm and reporting guidelines for HCC, the vocabulary between these two modalities and scoring criteria differs significantly. For instance, ancillary imaging features (favouring malignancy) are only documented in MRI exams but never appear in US while echogenicity which is an important characterization of lesion is US, cannot be visualized in MRI. We followed three different strategies for the language model transfer – (1) direct transfer – US LM was used to create embedding of the MRI reports. The words which were present in MRI space but not in US, were replaced by zero vector; (2) fine-tuning US LM on the MRI reports – US model fine-tuned with pre-processed text from EUH MRI reports since the language in US and MR reports are not drastically different, even when obtained from different hospitals; (3) fine-tuning Google News LM on the MRI reports – LM trained on 3 millions Google News articles fine-tuned with the pre-processed text from EUH MRI reports.

Mathematically, our objective in fine-tuning is that given the source US domain *D_US_* and the corresponding task of vectorization of words *T_US_*, learn the probability distribution *P*(*Y_MR_*|*X_MR_*) in the targeted MR domain *D_MR_* with the knowledge learnt from *D_US_* and *T_US_* where *Y_MR_* is the vector representation of words present in the MR vocabulary and *X_MR_* is the EUH MR reports. Note that even if both reports are from abdomen and liver disease mostly, the condition between the source *D_US_* and targeted *D_MR_* domain varies in several ways – (1) words used in the US and MRI domain are not same, i.e. *X_US_* = *X_MR_*; (2) marginal probability is distribution is also different between US and MRI domain, i.e. *P*(*X_MR_*) = *P*(*X_US_*).

We hypothesized that fine-tuning an existing US pre-trained language model trained on a large US dataset by continuously training it (i.e. running back-propagation) on the smaller MRI dataset should provide a more semantically meaningful network than training on the smaller dataset from scratch . Since our limited number of MRI reports did not allow us to completely retrain the model, we fine-tuned the US LM by freezing the weights for the words that were present in the US report corpus from Stanford. Using this approach, we only learnt the new words’ representation from the EUH MRI report corpus which in theory is a better solution than fine-tuning a LM trained on Google News where the vocabulary between Google News and MRI have limited overlap. We learnt the vocabulary from the EUH MRI reports by setting a minimum word frequency threshold. Thereafter, we merged an input-hidden weight matrix loaded from the US trained model, where it intersected with the current vocabulary. No extra words were added to the MRI vocabulary, but intersecting words adopted the US weights, and non-intersecting words were left alone to be solely learnt from the MRI corpus.

### Train discriminative model

For training the discriminative model to classify the reports, the liver section embedding was created by averaging the word vectors generated via by the models. According to earlier studies with generic text handling^19^ and also from our previous experience with radiology report classification tasks^13,14,15^, averaging the embeddings of words in a sentence/paragraph has been proven to be successful to obtain embeddings, where each section vector is computed as: 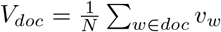, where *v_w_* is vector representation of the word *w, N* is the total number of word present in the document *doc* and *V_doc_* is the vector representation of the report. The dense and relatively lower dimensional numerical representation of the texts *V_doc_* is used as a feature vector to efficiently train a supervised classifier that can leverage the labeled dataset to learn the difference between benign and malignant reports. RandomForest model^20^, a meta estimator that fits a number of decision tree classifiers on various sub-samples of the dataset and then aggregates voting from the weak learners to improve the predictive accuracy and control over-fitting, was selected as the discriminative classifier. In all cases, we applied RandomForest model with default hyperparameters (number of estimators = 100, minimum sample split = 5).

### Statistical evaluation

We trained the machine learning models on the EUH structured MRI reports *(Training)* and tested the performance on the EUH historic free-text test set *(Test 1)* as well as on a subset of structured reports *(Test_2_)*. Among 944

EUH structured reports with ground truth label (Figure 2.a), we randomly selected 20% of the reports as our test dataset (189 reports) and the remaining reports (755 reports) were used to train our machine-learning model. We also evaluated our model which had been trained only on the structured reports, on a subset of randomly selected free-text reports (112 reports) documented without any structured reporting template annotated manually (Figure 2.b). The distribution of the targeted labels in our *Training, Test_1_* and *Test_2_* is shown in Table 2. In order to present a straight comparison between direct transfer of the US model (US language model) and fine-tuned models on the MRI cohort, we evaluated the models using the same discriminative classifier on both sets of structured *(Test 1)* and free-text reports (Test_2_).

**Table 2:**
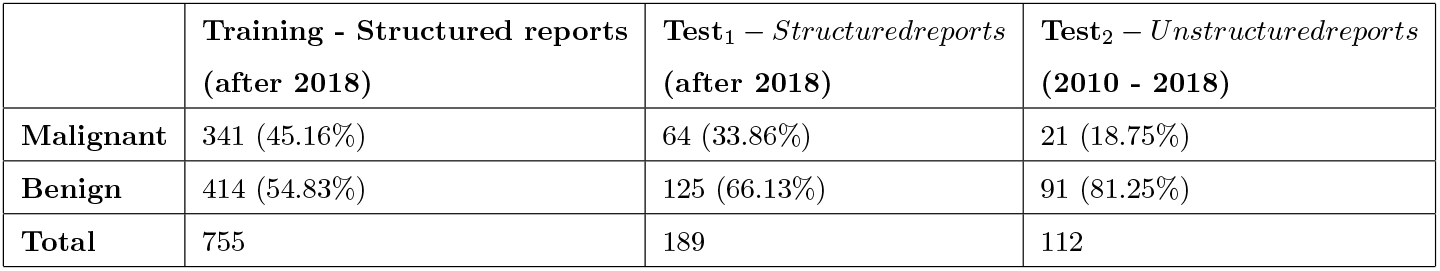
Distribution of targeted labels in training and testing set. Number of reports and % within each set has been reported.

|  | **Training - Structured reports**<br>**(after 2018)** | **Test<sub>1</sub> – Structured reports**<br>**(after 2018)** | **Test<sub>2</sub> – Unstructured reports**<br>**(2010 - 2018)** |
| --- | --- | --- | --- |
| **Malignant** | 341 (45.16%) | 64 (33.86%) | 21 (18.75%) |
| **Benign** | 414 (54.83%) | 125 (66.13%) | 91 (81.25%) |
| **Total** | 755 | 189 | 112 |

## Result

Interrater reliability was estimated as Cohen’s Kappa Score and the raters were highly consistent (kappa score 0.94) for identifying benign versus malignant cases by reading only the reports. A senior radiologist resolved all conflicting cases manually for preparing the ground truth labels. Table 3 presents the comparative performance in terms of class-wise precision, recall and f1-score along with number of evaluated cases.

**Table 3:** Performance of the NLP models on *Testi* and *Test2‘-* ‘US language model’ – direct transfer of the US model, ‘Fine-Tuned model GoogleNews Language model’ – LM trained on google news and fine-tuned on MRI, and ‘Fine-Tuned model US Language model’ – LM trained on US reports and fine-tuned on MRI,

|  | Report document with template |  |  |  | Report document without template |  |  |  |
| --- | --- | --- | --- | --- | --- | --- | --- | --- |
|  | Precision | Recall | f1-score | No. of cases | Precision | Recall | f1-score | No. of cases |
|  | US language model |  |  |  |  |  |  |  |
| *Malignant* | 0.91 | 0.92 | 0.91 | 64 | 0.68 | 0.62 | 0.65 | 21 |
| *Benign* | 0.96 | 0.95 | 0.96 | 125 | 0.91 | 0.93 | 0.92 | 91 |
|  | Fine-Tuned GoogleNews language model |  |  |  |  |  |  |  |
| *Malignant* | 0.87 | 0.91 | 0.89 | 64 | 0.65 | 0.62 | 0.63 | 21 |
| *Benign* | 0.95 | 0.93 | 0.94 | 125 | 0.91 | 0.92 | 0.92 | 91 |
|  | Fine-Tuned US language model |  |  |  |  |  |  |  |
| *Malignant* | **0.98** | **0.92** | **0.95** | 64 | **0.78** | **0.67** | **0.72** | 21 |
| *Benign* | **0.96** | **0.99** | **0.98** | 125 | **0.93** | **0.96** | **0.94** | 91 |

Figure 3 presents the visualization of dimension reduced language space (PCA reduced dimension from 300 → 2) learnt from only US reports (a), fined-tuned US language space on the MRI report corpus (b), and also the overlapping language spaces (c). Each point in the spaces represents unique word in the vocabulary. As seen from the overlapping language spaces (zoomed representation Figure 3.d), fine-tuning strategy on a small subset of MRI reports was not only able to learn the new words in the MRI domain but was also able to preserve the overall semantics of the US space and learnt correlation between words in two different domains. For instance, ‘hyperintense’ is a word from MRI space but one of its synonyms is ‘bright’ learnt from the US corpus.

**Figure 3:**
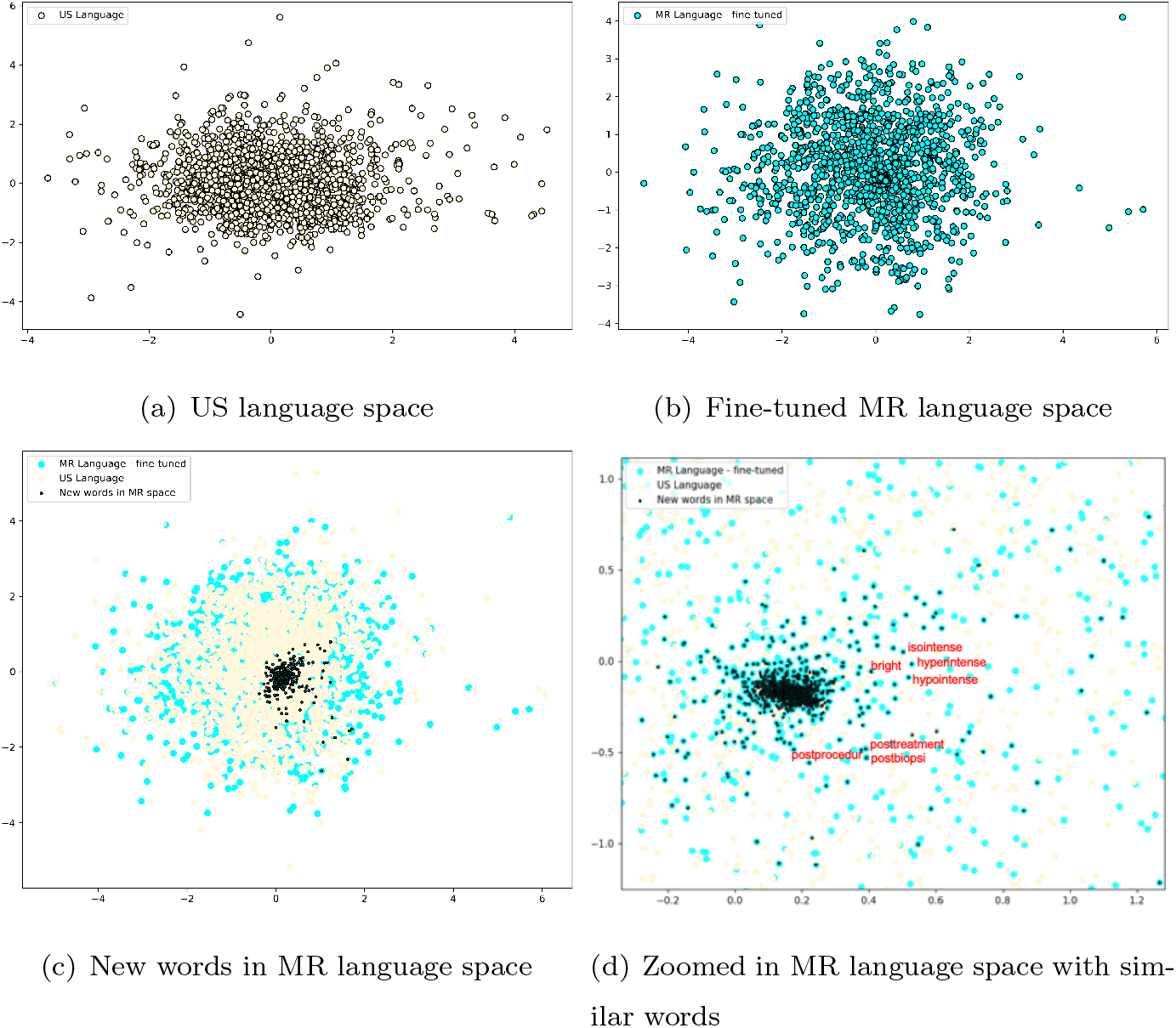
Represent distinct language spaces after dimension reduction using TSNE

The classifier running on report vectors generated by the fine-tuned US LM outperformed the directly transferred LM on all cases. When tested on the LI-RADS structured reports, the fine-tuned US model achieved f1-score of 0.95 for identifying malignant cases while the model with direct transfer scored 0.91. For benign cases, the fine-tuned US model achieved 0.98 f1-score while the model with direct transfer scored 0.96. The performance on structured reports improved with fine-tuning the language space but direct transfer model also resulted in high accuracy for identifying both benign and malignant cases.

Fine-tuned US LM outperformed fine-tuned Google news LM on the benign and malignant categories. On the structured reports, the fine-tuned Google news model achieved 0.89 f1-score for identifying malignant cases while the model with direct transfer scored 0.95. For benign cases, the fine-tuned google news model achieved 0.94 f1-score while the fine-tuned US model with direct transfer scored 0.98.

The major improvement while fine-tuning the language space is observed during labeling of the free-text dataset which is a more challenging task for a classifier that has only been trained on the structured reports. For example, while labeling malignant cases, the classifier with fine-tuned US LM improved the f1-score from 0.65 to 0.72 by correctly classifying non-trivial cases where the other model failed, e.g. cases with no HCC but other abnormality. The performance on benign cases also improved with fine-tuning the US LM from 0.92 to 0.94. Interestingly, fine-tuning the google news LM only achieved 0.63 f1-score on malignant cases and performance stayed the same as of direct transfer of the US model on the benign cases. Our results demonstrate that fine-tuning LM on similar domain helps the LM to converge fast on a small dataset.

## Discussion

The study reports a successful transfer of language models from one domain (US) to a similar domain (MRI) and compares it to not performing adaptation and to adapting a more generic model (trained on Google News). The core novelty of this whole pipeline is that only a limited domain-specific corpus is required to train the word-embedding model while performing the LM transfer. We showed that fine-tuning of the word-embedding model with similar domain adaptation (US → MRI) provides more opportunity for semantic knowledge preservation for down-steam classification tasks compared to fine-tuning a completely different domain model (Google news → MRI radiology reports).

Our study provides a way to train language space model with limited number of reports by using cross-domain transfer. Our proposed pipeline automatically extracted binary labels (benign/malignant) for MRI imaging dataset for HCC screening with high accuracy. Our proposed framework only need a small subset of labeled MR exams to train the discriminative model on top of the dense vector embedding to generate a large dataset of labeled MR exams. The labeled MR studies may be used for AI development and training, and such automated NLP methods will rapidly reduce the manual workload for creating labeled imaging datasets from hospital databases.

This study lacks experimentation with the sensitivity for word order that limits the ability of learning long term and rotated scope of negex term. However, thanks to the semantic mapping and word occurrence analysis in out study, the adjacent negex terms are concatenated with the targeted entity in the preprocessed text before Word2Vec training, e.g. ‘negex_abnormal’. As a result, the model had the opportunity to learn the representation of the entity and the adjacent negation of the entity without explicitly considering the sentence boundary. In future, we plan to collect a larger multi-institutional radiology reports corpus and perform experimentation with advanced Bert (Bidirectional Encoder Representations from Transformer) sequential embedding model.

## Data Availability

Available upon reasonable request

## Acknowledgement

The study is supported by funding received from Winship Cancer Institute, and partially supported by ‘Stanford NLP collaboration’ funding.

